# POST-ACUTE SEQUELAE AND ADAPTIVE IMMUNE RESPONSES IN PEOPLE LIVING WITH HIV RECOVERING FROM SARS-COV-2 INFECTION

**DOI:** 10.1101/2022.02.10.22270471

**Authors:** Michael J. Peluso, Matthew A. Spinelli, Tyler-Marie Deveau, Carrie A. Forman, Sadie E. Munter, Sujata Mathur, Alex F. Tang, Scott Lu, Sarah A. Goldberg, Mireya I. Arreguin, Rebecca Hoh, Viva Tai, Jessica Y. Chen, Enrique O. Martinez, Ahmed Chenna, John W. Winslow, Christos J. Petropoulos, Alessandro Sette, Daniella Weiskopf, Nitasha Kumar, Kara L. Lynch, Peter W. Hunt, Matthew S. Durstenfeld, Priscilla Y. Hsue, J. Daniel Kelly, Jeffrey N. Martin, David V. Glidden, Monica Gandhi, Steven G. Deeks, Rachel L. Rutishauser, Timothy J. Henrich

## Abstract

**Background:** Limited data are available on the long-term clinical and immunologic consequences of SARS-CoV-2 infection in people with HIV (PWH).

**Methods:** We measured SARS-CoV-2 specific humoral and cellular responses in people with and without HIV recovering from COVID-19 (n=39 and n=43, respectively) using binding antibody, surrogate virus neutralization, intracellular cytokine staining, and inflammatory marker assays. We identified individuals experiencing post-acute sequelae of SARS-CoV-2 infection (PASC) and evaluated immunologic parameters. We used linear regression and generalized linear models to examine differences by HIV status in the magnitude of inflammatory and virus-specific antibody and T cell responses, as well as differences in the prevalence of PASC.

**Results:** Among PWH, we found broadly similar SARS-CoV-2-specific antibody and T cell responses as compared with a well-matched group of HIV-negative individuals. PWH had 70% lower relative levels of SARS-CoV-2 specific memory CD8+ T cells (p=0.007) and 53% higher relative levels of PD-1+ SARS-CoV-2 specific CD4+ T cells (p=0.007). Higher CD4/CD8 ratio was associated with lower PD-1 expression on SARS-CoV-2 specific CD8+ T cells (0.34-fold effect, p=0.02). HIV status was strongly associated with PASC (odds ratio 4.01, p=0.008), and levels of certain inflammatory markers (IL-6, TNF-alpha, and IP-10) were associated with persistent symptoms.

**Conclusions:** We identified potentially important differences in SARS-CoV-2 specific CD4+ and CD8+ T cells in PWH and HIV-negative participants that might have implications for long-term immunity conferred by natural infection. HIV status strongly predicted the presence of PASC. Larger and more detailed studies of PASC in PWH are urgently needed.

## BACKGROUND

The intersection between the SARS-CoV-2 and HIV epidemics has gained increased attention [1]. While studies conducted early in the course of the pandemic did not show differences in outcomes of acute COVID-19 associated with HIV status [2,3], larger carefully designed studies show that people with HIV (PWH) are at higher risk for adverse outcomes [4,5]. Data on SARS-CoV-2-specific adaptive immune responses in PWH compared with HIV-negative individuals remain sparse, however, with one study showing less robust immune responses among PWH [6] but another [7] suggesting similar responses. Furthermore, there is growing recognition of the clinical burden of post-acute sequelae of SARS-CoV-2 infection (PASC, including “long COVID”) [8], but this condition remains poorly understood, especially in PWH.

While the mechanisms underlying PASC have yet to be fully delineated, early studies have suggested that a combination of factors might contribute [9–14]. Higher prevalence of certain socioeconomic factors and medical comorbidities among PWH, along with differences in immune responses to SARS-CoV-2 [6,7] and persistent inflammation and immune dysregulation in the presence of antiretroviral therapy (ART) [15–18], may make PWH selectively vulnerable to developing persistent COVID-19-attributed symptoms following SARS-CoV-2 infection. For these reasons, examination of PASC in PWH is urgently needed.

Here, we sought to test the hypothesis that PASC would be more prevalent in a cohort of PWH with a history of COVID-19 prior to vaccination and that these individuals would have reduced SARS-CoV-2 specific immune responses in comparison to a similar group of HIV-negative individuals recovering from COVID-19.

## METHODS

### Informed consent

The study was approved by the Institutional Review Board at the University of California, San Francisco (UCSF). All participants provided written informed consent.

### Participants

Volunteers were enrolled in the Long-term Impact of Infection with Novel Coronavirus (LIINC) COVID-19 recovery cohort at UCSF (NCT04362150). Study procedures are described in detail elsewhere [19,20]. Briefly, participants with a prior positive nucleic acid amplification test confirming SARS-CoV-2 infection were enrolled 21 or more days following symptom onset. Recruitment occurred through a combination of self-and clinician referrals; all PWH testing positive for SARS-CoV-2 at two UCSF-based HIV clinics were notified of the study and invited to participate. At each study visit, participants completed a structured interview in which they provided clinical data regarding prior and current COVID-19-attributed symptoms, medical history, and quality of life.

We selected all PWH who enrolled prior to the receipt of a SARS-CoV-2 vaccine (n=39) and compared them with a randomly selected group of HIV-negative individuals (n=43) with a similar distribution of key variables reported to be important in determination of the SARS-CoV-2 immune responses in our cohort [9,21–23]: age, sex, COVID-19 hospitalization, and time since infection. Participants were assessed at the time point closest to 16 weeks post-infection (median 112 days [IQR: 91-129]).

### Clinical assessment of PASC

Our methodology for assessing PASC in the cohort is described in detail elsewhere [20]. Briefly, responses to questions regarding the presence of 32 somatic symptoms derived from the U.S. Centers for Disease Control (CDC) list of COVID-19 symptoms (https://www.cdc.gov/coronavirus/2019-ncov/symptoms-testing/symptoms.html) and the Patient Health Questionnaire Somatic Symptom Scale [24] were characterized at each study visit. Symptoms are only considered as related to COVID-19 if they are new or worsened since the time of initial SARS-CoV-2 infection; stable, chronic symptoms that pre-dated SARS-CoV-2 infection are not included in the PASC definition.

For the primary analysis, we defined PASC as any COVID-19 attributed symptom that was present during a study visit >6 weeks following SARS-CoV-2 infection following consensus definitions [25,26]. We conducted further sensitivity analyses, comparing those with 3 or more symptoms to those with fewer than 3 symptoms or with no symptoms.

### Biospecimen isolation

Peripheral blood mononuclear cells (PBMCs), plasma and serum were obtained from participants and cryopreserved prior to testing. PBMCs were isolated using Ficol-Paque in SeptMate Tubes. PBMCs were frozen in heat-inactivated FBS and 10% dimethyl sulfoxide (DMSO) and stored in liquid nitrogen.

### Antibody assays

Virus-specific antibody responses were measured using the Pylon COVID-19 total antibody assay (ET Health) and a validated surrogate virus neutralization test (sVNT) [27]. The sVNT measures competitive inhibition of the interaction of the original SARS-CoV-2 receptor binding domain (RBD) and angiotensin-converting enzyme 2 (ACE-2). The lower limit of detection was a sVNT reciprocal titer <10 and anti-RBD IgG <10 relative fluorescence units.

### T cell responses by Intracellular Cytokine Staining (ICS)

We performed ICS as previously described [22] with the following modifications. Two previously described peptide megapools containing SARS-CoV-2-derived peptides experimentally determined to be recognized by CD4+ or CD8+ T cells were used for 18-hour peptide stimulations (1 μg/ml/peptide; CD4-E with 280 and CD8-E with 454 T cell epitopes) [28]. Additional detail and a complete list of antibodies are listed in Supplementary Methods. All samples were acquired on a BD LSR-II analyzer and analyzed with FlowJo X software. The flow cytometry gating strategy is shown in Supplementary Figure 1.

### Markers of inflammation

In a subset of participants with additional specimens available, the fully automated HD-X Simoa platform was used to measure biomarkers in blood plasma including monocyte chemoattractant protein 1 (MCP-1), Cytokine 3-PlexA (IL-6, IL-10, TNFα), interferon gamma-induced protein-10 (IP-10), and interferon-gamma (IFNγ). Samples were run blinded. Assay performance was consistent with the manufacturer’s specifications.

### Statistical methods

We used descriptive statistics to characterize the cohort. For the comparison of humoral responses, we first log-transformed sVNT and total antibody values to satisfy assumptions of a normal distribution. We used linear regression models to examine differences in magnitude of humoral models by HIV status, adjusting for days since onset of known SARS-CoV-2 infection, age, sex at birth, history of hospitalization for SARS-CoV-2 infection, and the primary indicator, HIV status. Differences and 95% confidence intervals (CI) are presented as fold-changes in the geometric mean. Among PWH, CD4/CD8 ratio was then added to the model as a covariate to examine if it was associated with magnitude of responses among PWH. To examine percent differences in cellular immune responses by HIV status, we used generalized linear models from the binomial distribution with bootstrapped standard errors and adjusted for the same potential confounders as above. To examine differences in relative mean inflammatory markers by HIV-status, we fit linear regression models following log-transformation of the individual inflammatory markers. To examine differences in the experience of PASC by HIV status, we used logistic regression adjusting for the same potential confounders.

## RESULTS

### Participants

All participants were diagnosed with COVID-19 by nucleic acid amplification (NAA) testing between March and December 2020 (pre-Delta and Omicron) and none had received a SARS-CoV-2 vaccine prior to specimen collection. The groups were similar in terms of age, sex, and race/ethnicity (Table 1). The vast majority of PWH in the study were men, reflecting the demographics of the local HIV epidemic [29]. While most participants were generally healthy, those with HIV more commonly reported concurrent medical comorbidities including heart and lung disease. The majority of participants were managed as outpatients during their acute COVID-19 illness. All previously hospitalized participants required supplemental oxygenation and one in each group required mechanical ventilation. No participant received SARS-CoV-2-specific treatment during their acute illness.

**Table 1.**
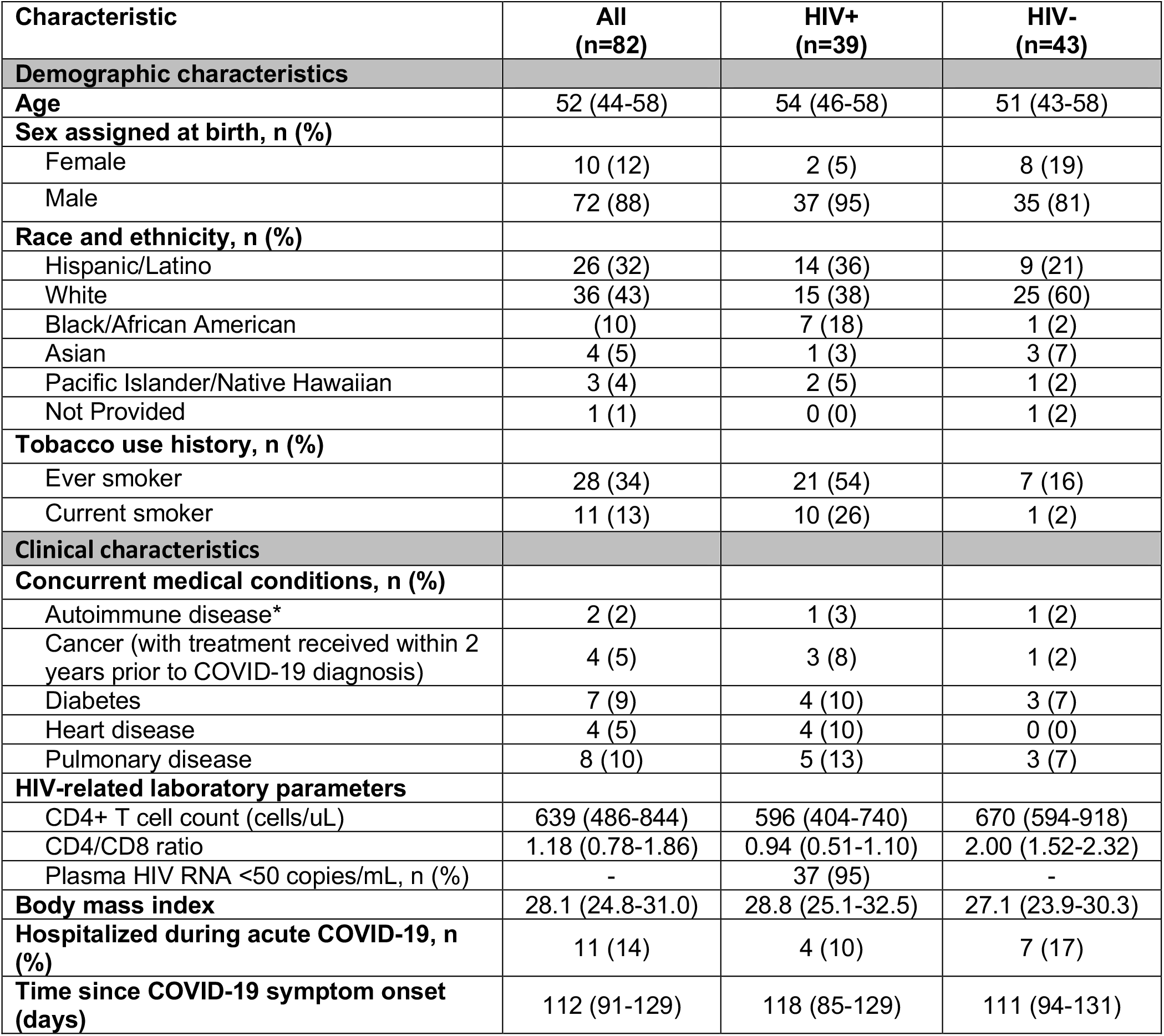
Characteristics of study cohort. Values reported as median (IQR) unless otherwise specified. IQR, interquartile range. Note: Autoimmune disease reported as one case of hypothyroidism and one case of Raynaud’s disease. Cancer reported as prostate cancer controlled with hormone therapy, ocular melanoma, treated Kaposi sarcoma, and resected renal cancer. Plasma HIV RNA values were >50 in two participants (87 copies/mL and 28,118 copies/mL). All hospitalized participants in each group required oxygen support and one hospitalized participant in each group required mechanical ventilation. None reported receiving SARS-CoV-2-targeted therapy or steroids during the hospitalization.

| Characteristic | All<br>(n=82) | HIV+<br>(n=39) | HIV-<br>(n=43) |
| --- | --- | --- | --- |
| **Demographic characteristics** |  |  |  |
| **Age** | 52 (44-58) | 54 (46-58) | 51 (43-58) |
| **Sex assigned at birth, n (%)** |  |  |  |
| Female | 10 (12) | 2 (5) | 8 (19) |
| Male | 72 (88) | 37 (95) | 35 (81) |
| **Race and ethnicity, n (%)** |  |  |  |
| Hispanic/Latino | 26 (32) | 14 (36) | 9 (21) |
| White | 36 (43) | 15 (38) | 25 (60) |
| Black/African American | (10) | 7 (18) | 1 (2) |
| Asian | 4 (5) | 1 (3) | 3 (7) |
| Pacific Islander/Native Hawaiian | 3 (4) | 2 (5) | 1 (2) |
| Not Provided | 1 (1) | 0 (0) | 1 (2) |
| **Tobacco use history, n (%)** |  |  |  |
| Ever smoker | 28 (34) | 21 (54) | 7 (16) |
| Current smoker | 11 (13) | 10 (26) | 1 (2) |
| **Clinical characteristics** |  |  |  |
| **Concurrent medical conditions, n (%)** |  |  |  |
| Autoimmune disease* | 2 (2) | 1 (3) | 1 (2) |
| Cancer (with treatment received within 2 years prior to COVID-19 diagnosis) | 4 (5) | 3 (8) | 1 (2) |
| Diabetes | 7 (9) | 4 (10) | 3 (7) |
| Heart disease | 4 (5) | 4 (10) | 0 (0) |
| Pulmonary disease | 8 (10) | 5 (13) | 3 (7) |
| **HIV-related laboratory parameters** |  |  |  |
| CD4+ T cell count (cells/uL) | 639 (486-844) | 596 (404-740) | 670 (594-918) |
| CD4/CD8 ratio | 1.18 (0.78-1.86) | 0.94 (0.51-1.10) | 2.00 (1.52-2.32) |
| Plasma HIV RNA <50 copies/mL, n (%) | - | 37 (95) | - |
| **Body mass index** | 28.1 (24.8-31.0) | 28.8 (25.1-32.5) | 27.1 (23.9-30.3) |
| **Hospitalized during acute COVID-19, n (%)** | 11 (14) | 4 (10) | 7 (17) |
| **Time since COVID-19 symptom onset (days)** | 112 (91-129) | 118 (85-129) | 111 (94-131) |

The median time between COVID-19 symptom onset and research assessment was 117 (85-128) days for PWH and 111 (94-131) days for HIV-negative individuals (p=0.58; Table 2). Initial illness severity was similar between both groups (13% vs 17% hospitalized, p=0.41) Participants reported a wide array of symptoms attributed to COVID-19 at the time of PASC assessment (Table 2). There were similarities in post-acute symptoms reported among both groups, but a larger proportion of PWH reported certain symptoms such as fatigue, gastrointestinal and certain neurocognitive symptoms, and issues with sleep.

**Table 2.** Symptoms reported at late follow-up among those with PASC. Values reported as n (%). Participants were systematically asked about 32 individual symptoms at the late follow-up visit, which took place a median of 124 days from initial COVID-19 symptom onset.

| Symptoms reported at late follow-up | All<br>(n=82) | HIV+<br>(n=39) | HIV-<br>(n=43) |
| --- | --- | --- | --- |
| Constitutional |  |  |  |
| *Fatigue* | 26 (32) | 16 (42) | 10 (23) |
| *Subjective fever* | 1 (1) | 1 (3) | 0 (0) |
| *Chills* | 2 (2) | 2 (5) | 0 (0) |
| *Objective fever* | 0 (0) | 0 (0) | 0 (0) |
| Upper Respiratory |  |  |  |
| *Rhinorrhea* | 13 (16) | 5 (13) | 8 (19) |
| *Sore throat* | 2 (2) | 0 (0) | 2 (5) |
| Cardiopulmonary |  |  |  |
| *Cough* | 7 (9) | 3 (8) | 4 (9) |
| *Shortness of breath* | 20 (25) | 11 (29) | 9 (21) |
| *Chest pain* | 7 (9) | 3 (8) | 4 (9) |
| *Palpitations* | 10 (12) | 4 (11) | 6 (14) |
| *Fainting* | 0 (0) | 0 (0) | 0 (0) |
| Gastrointestinal |  |  |  |
| *Diarrhea* | 7 (9) | 5 (13) | 2 (5) |
| *Nausea* | 9 (11) | 5 (13) | 4 (9) |
| *Loss of appetite* | 4 (5) | 3 (8) | 1 (2) |
| *Abdominal pain* | 9 (11) | 5 (13) | 4 (9) |
| *Vomiting* | 1 (1) | 0 (0) | 1 (2) |
| *Constipation* | 0 (0) | 0 (0) | 0 (0) |
| Genitourinary |  |  |  |
| *Menstrual cramps* | 0 (0) | 0 (0) | 0 (0) |
| *Dyspareunia* | 0 (0) | 0 (0) | 0 (0) |
| Rash | 8 (10) | 7 (18) | 1 (2) |
| Musculoskeletal |  |  |  |
| *Myalgia* | 14 (17) | 9 (24) | 5 (12) |
| *Back pain* | 5 (6) | 5 (13) | 0 (0) |
| *Joint pain* | 11 (14) | 8 (21) | 3 (7) |
| Anosmia/Dysgeusia | 14 (17) |  |  |
| Neurologic |  |  |  |
| *Headache* | 14 (17) | 9 (24) | 5 (12) |
| *Concentration problems* | 24 (30) | 16 (42) | 8 (19) |
| *Dizziness* | 11 (14) | 5 (13) | 6 (14) |
| *Balance problems* | 9 (11) | 6 (16) | 3 (7) |
| *Neuropathy* | 9 (11) | 7 (18) | 2 (5) |
| *Vision problems* | 11 (14) | 8 (21) | 3 (7) |
| *Parosmia* | 2 (2) | 0 (0) | 2 (5) |
| Trouble Sleeping | 20 (24) | 13 (34) | 7 (16) |

### SARS-CoV-2-specific antibody responses

Antibody responses were similar between people with and without HIV infection. In models incorporating HIV status, days since infection, age, sex, and prior hospitalization during SARS-CoV-2 infection (yes vs. no), HIV status was not a predictor of humoral responses as measured by SARS-CoV-2-specific antibody binding (1.31-fold higher; 95%CI: 0.70-2.46; p=0.40; Fig. 1a) and surrogate virus neutralization testing (1.01-fold higher; 95% CI: 0.63-1.63; p=0.95; Fig. 1b). Hospitalization during acute COVID-19 was associated with 4.29-fold higher geometric mean titers of binding antibodies (95% CI:1.74-10.57; p=0.002) and 2.57-fold higher titers of surrogate viral neutralization testing (95% CI: 1.33-4.98; p=0.005).

**Figure 1.**
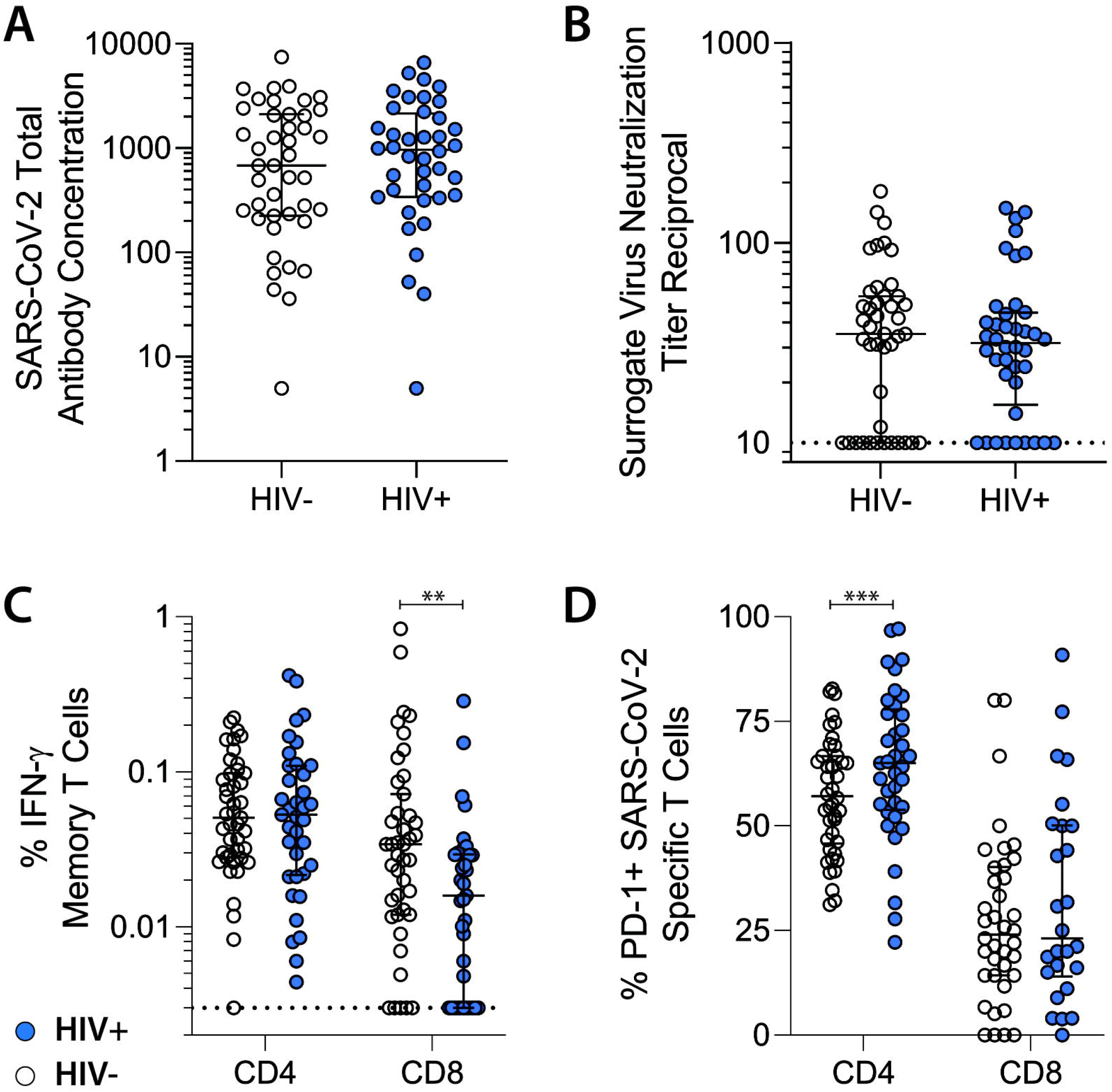
SARS-CoV-2 specific immunologic parameters among people with and without HIV recovering from COVID-19. A. Total SARS-CoV-2 antibodies. B. Surrogate SARS-CoV-2 neutralization titres. C. Frequency of IFNγ+ SARS-CoV-2 specific memory CD4+ and CD8+ T cells. D. PD-1 expression on SARS-CoV-2-specific CD4+ and CD8+ T cells. Bars represent median and interquartile ranges. **p<0.01; *** p<0.001 in covariate adjusted linear regression modeling.

### SARS-CoV-2 specific T cell responses

In models incorporating days since infection, age, sex, and prior hospitalization during SARS-CoV-2 infection, HIV status was not a predictor of the magnitude of interferon gamma (IFNL)-producing SARS-CoV-2-specific memory CD4+ T cell responses using the SARS-CoV-2 CD4+ T cell epitope-focused megapool (1.14-fold higher; 95%CI: 0.76-1.71; p=0.55; median: 0.070% vs. 0.068%; Fig. 1c). However, PWH had 70% lower relative levels of SARS-CoV-2-specific memory CD8+ T cells using a CD8+ T cell epitope-focused megapool (0.30-fold 95% CI: 0.13-0.72; p=0.007; median: 0.016% vs 0.034%; Fig. 1c).

PWH exhibited higher levels of PD-1 expression on SARS-CoV-2-specific memory CD4+ T cells in adjusted regression analyses (1.18-fold higher; 95% CI: 1.07-1.30; p=0.001; median: 65% vs. 57.1%; Fig. 1d), but no significant differences in PD-1 expression on SARS-CoV-2 specific CD8+ T cells (1.21-fold higher; 95% CI: 0.83-1.76; p=0.33; median: 25.0% vs. 24.1%; Fig. 1d).

### Effect of CD4/CD8 ratio

We next examined whether the CD4/CD8 ratio was associated with adaptive immune responses after infection with SARS-CoV-2 among PWH using adjusted regression modeling as above with the addition of CD4/CD8 ratio as a covariate. The ratio was not predictive of binding antibody levels (p=0.30) or surrogate virus neutralization (p=0.61). Higher ratios were associated with 67% lower overall frequency of SARS-CoV-2 specific CD4+ T cells (0.33-fold; 95% CI: 0.19-0.97; p=0.04) and 36% lower SARS-CoV-2 specific PD-1 expression (0.64-fold 95% CI:0.57-0.97; p=0.03). Notably, higher CD4/CD8 ratios were also associated with 66% lower PD-1 expression on SARS-CoV-2 specific CD8+ T cells (0.34-fold 95% CI: 0.13-0.87; p=0.02). There was a nonsignificant trend toward a similar finding among HIV-negative individuals (0.70-fold; 95% CI: 0.47-1.04; p=0.08).

### Relationship between antibody and T cell immune responses

Similar to our prior work [21], we observed strong non-parametric correlations between the binding (r=0.33, p=0.008) and surrogate viral neutralization response (r=0.33, p=0.007) and between binding and surrogate viral neutralization responses and SARS-CoV-2 specific CD4+ T cells (r=0.41, p<0.001 and r=0.42, p<0.001, respectively).

### Markers of systemic inflammation

PWH had higher levels of plasma markers of inflammation compared with HIV-negative comparators. Among PWH, mean IL-6 levels were 1.55-fold higher (95% CI: 1.06-2.26; p=0.02), mean IP-10 levels were 1.31-fold higher (95% CI: 1.06-1.62; p=0.01), and TNFα levels were 1.26-fold higher (95% CI: 1.08-1.47; p=0.003) than in HIV-negative comparators.

### Post-acute sequelae

We found HIV status to be highly associated with the presence of PASC. In the primary analysis, PWH had 4.01-fold higher odds of reporting PASC (95% CI: 1.45-11.1; p=0.008; 82.8% vs. 54.4%), in a model controlling for time since infection, hospitalization, and age. This relationship was maintained when defining PASC as 3 or more symptoms in comparison to fewer than 3 symptoms (adjusted odds ratio (AOR) 2.72; 1.08-6.88; p=0.03; 59.8% vs. 33.6%). PWH reported more somatic symptoms overall (median 3 [IQR 1-6] versus median 1 [IQR 0-5], p=0.02), and in a model including time since infection, hospitalization, sex, and age, those with HIV had a 1.91-fold higher number of individual PASC symptoms than those without HIV (p=0.02).

Antibody and T cell responses did not correlate with PASC within the cohort (Fig. 2a-d). In models adjusting for HIV status, higher PD-1 expression on total memory CD4+ T cells, but not memory CD8+ T cells, was independently predictive of lower odds of PASC (Fig. 2e-f). However, when stratifying by HIV status, this finding appeared to be driven by differences in overall PD-1 expression between the HIV-positive and -negative groups, in which no significant difference between those with and without PASC was identified. Furthermore, there appeared to be a trend toward higher PD-1 expression among those with PASC in stratified analyses (Fig. 2e-f). There was no relationship between PD-1 expression on SARS-CoV-2-specific CD4+ or CD8+ T cells and PASC (AOR 0.23; 95% CI: 0.01-6.31; p=0.39 and AOR 0.20; 0.01-3.50; p=0.27 respectively; Fig. 2g-h).

**Figure 2.**
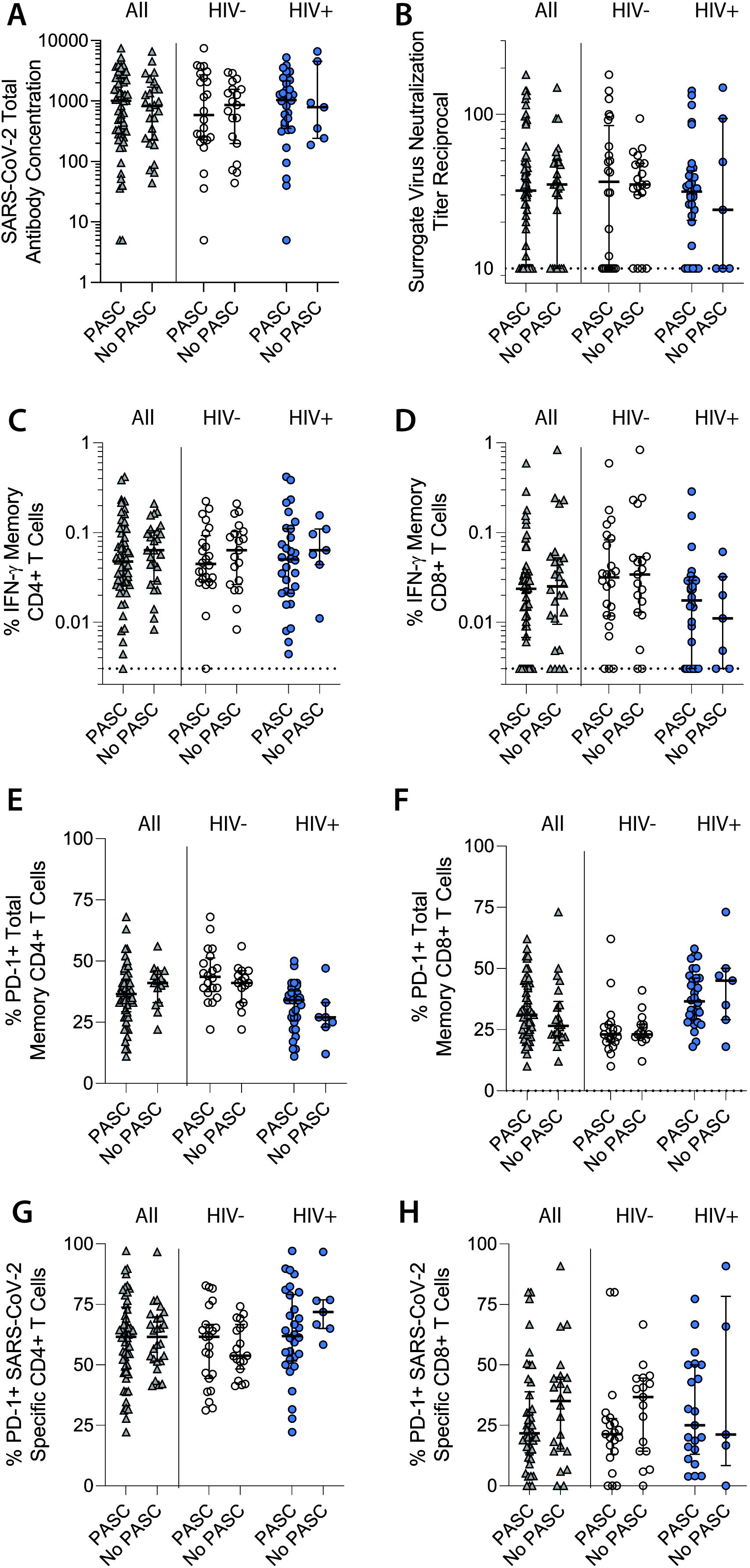
SARS-CoV-2 specific immune responses among people with and without PASC, stratified by HIV status. A. Total SARS-CoV-2 antibodies. B. Surrogate viral neutralization titers. C. SARS-CoV-2-specific CD4+ T cells. D. SARS-CoV-2-specific CD8+ T cells. E. PD-1 expression on total memory CD4+ T cells. F. PD-1 expression on total memory CD8+ T cells. G. PD-1 expression on SARS-CoV-2-specific memory CD4+ T cells. H. PD-1 expression on SARS-CoV-2-specific memory CD8+ T cells. Bars represent median and interquartile ranges.

We found that some inflammatory markers were associated with increased odds of PASC (Fig. 3a-e). After adjusting for HIV status, the odds of PASC in the entire study population increased 1.18-fold for each 10% increase in IP-10 (AOR 1.18; 95% CI:1.01-1.38; p=0.04); and 1.10-fold for each 10% increase in IL-6 (AOR 1.10; 95% CI: 1.01-1.21; p=0.04); while there was a trend in increased PASC with higher TNFα levels (AOR 1.19; 95% CI: 0.98-1.46; p=0.08). Other plasma markers of inflammation were not associated with experience of PASC, including IL-10 (p=0.61), interferon gamma (p=0.73), and MCP-1 (p=0.93).

**Figure 3.**
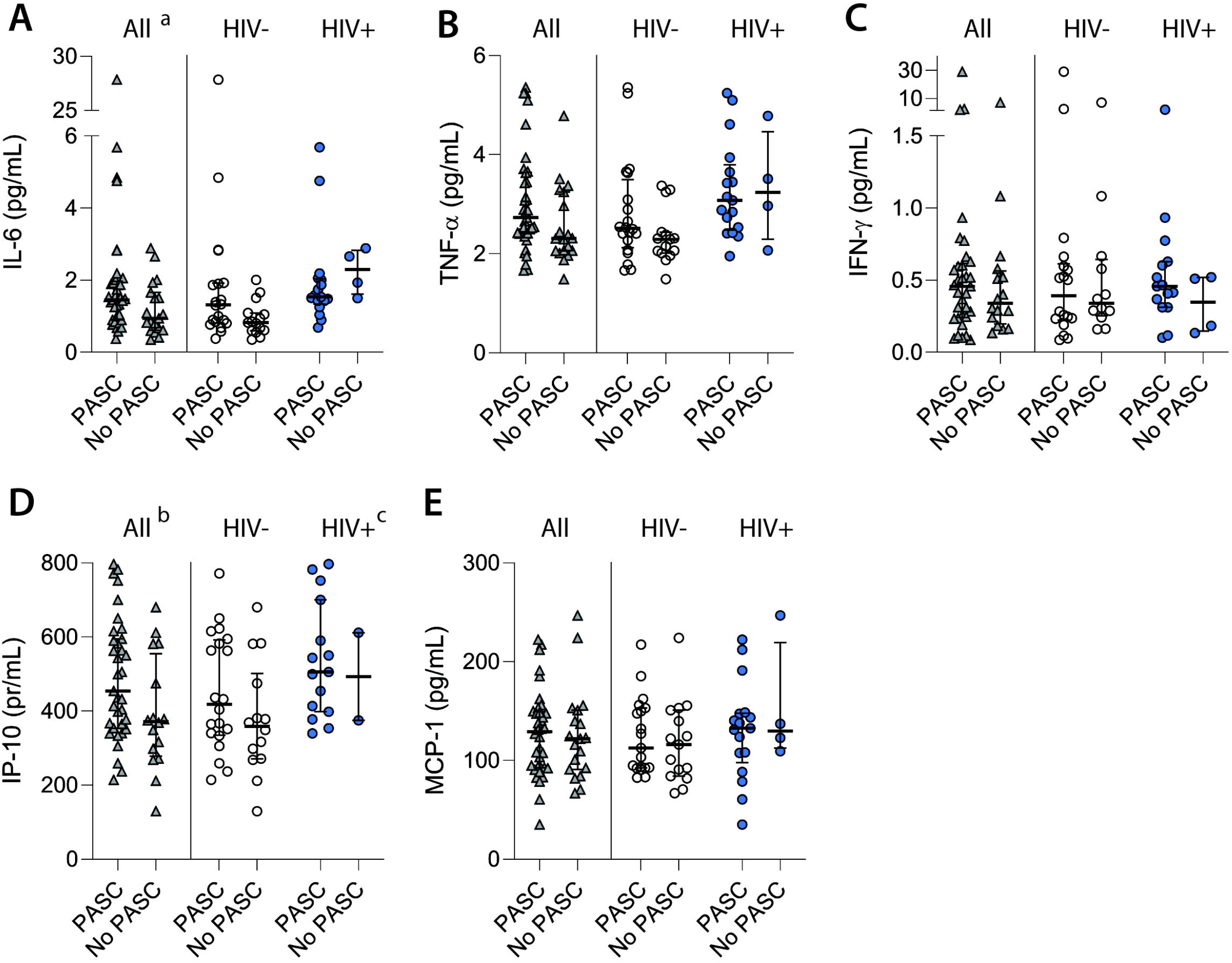
Plasma markers of immune activation among people with and without PASC, stratified by HIV status. A. Interleukin-6 levels. B. TNF-alpha levels. C. Interferon-gamma levels. D. IP-10 levels. E. MCP-1 levels. Bars represent median and interquartile ranges. ^a^The odds of PASC of the entire study population increased 1.18-fold for each 10% increase in IP-10 (AOR 1.18; 95% CI:1.01-1.38; p=0.04); and ^b^1.10-fold for each 10% increase in IL-6 (AOR 1.10; 95% CI: 1.01-1.21; p=0.04) from adjusted linear regression. ^c^Among PWH, there were increased odds of PASC with each 10% increase in IP-10 levels (AOR 1.06; 95% CI: 1.00-1.11; p=0.05).

Among PWH, there were increased odds of PASC with each 10% increase in IP-10 levels (AOR 1.06; 95% CI: 1.00-1.11; p=0.05), and a trend for increased PASC with higher TNFα levels (AOR 1.20 per 10% increase; 95% CI: 0.97-1.49; p=0.09), but not IL-6 (p=0.64). This analysis was limited by the relatively small number of individuals with HIV who reported full recovery.

## DISCUSSION

We observed that HIV status was strongly associated with PASC, raising concerns that this condition might be common among PWH recovering from COVID-19. Higher levels of inflammation were significantly associated with PASC. Finally, we observed potentially important differences in SARS-CoV-2-specific CD4+ and CD8+ T cells that might have implications for long-term immunity conferred by natural infection. This study adds to the limited published data examining SARS-CoV-2-specific immune responses in PWH and underscores the need for larger and more detailed studies of PASC in this population.

While there are massive efforts underway to understand the prevalence and pathophysiology of PASC, there are limited data among PWH [30]. One single-center study suggested that acute illness severity in PWH was associated with PASC with 44% of participants experiencing at least one symptom 30 days after recovery [31], however it did not find an association with CD4+ T cell count, viral load, demographics or comorbidities, and did not compare PWH with HIV-negative individuals. Another study suggested that HIV was one factor associated with higher risk of PASC among those requiring emergency department or hospital-based care [32], but the analysis was limited to 10 PWH and did not include biological measurements. While our cohort is not representative of all individuals with SARS-CoV-2 infection and cannot estimate the population-level prevalence of PASC, the observation that persistent SARS-CoV-2-attributed symptoms were highly prevalent in PWH and that the adjusted odds of PASC were four-fold as high as in well-matched HIV-negative comparators was striking. Large-scale epidemiologic studies in which HIV can be examined as a predictor of PASC are urgently needed.

PASC may be driven, at least in part, by residual or ongoing inflammation following SARS-CoV-2 infection [9,10]. ART-treated HIV is a chronic inflammatory condition associated with persistent immune activation and inflammation [15–18]. Further immune perturbations related to COVID-19 may therefore lead to a higher prevalence of PASC among PWH. Additional factors could also predispose PWH to PASC, such as autoimmunity [33], localized tissue inflammation [34], human herpesvirus reactivation [13], and microvascular dysfunction [14], among others. Other comorbidities including substance use and metabolic disorders may further contribute. Regardless of the mechanism, our observation is of importance because it suggests that PASC may be especially common in PWH and emphasizes the need for studies of PASC in this population.

Data comparing SARS-CoV-2-specific adaptive immune responses in people with and without HIV remain limited. Given the association between the presence of potent, durable SARS-CoV-2-specific immune responses and protection from disease upon re-exposure, it is critical to understand how HIV may modulate protective immunity. Furthermore, there is evidence that SARS-CoV-2 can cause chronic infection in certain immunocompromised individuals, including those with advanced HIV infection [35]. Since the beginning of the pandemic, there has been concern that PWH are less likely to develop and maintain protective immunity. While some studies suggested lower humoral responses in PWH [6], we and others [36–38] have not observed this.

Data on cellular immune responses to SARS-CoV-2 infection in PWH are even more limited. A single high-quality study has shown similar T cell responses between PWH on ART and HIV-negative individuals [7]. Our findings contribute three key observations regarding SARS-CoV-2-specific cellular immune responses in PWH. First, using peptide pools that include optimal SARS-CoV-2 epitopes spanning the proteome, we found that PWH had lower SARS-CoV-2 specific CD8+ T cell responses. This difference was previously observed in non-Spike-specific T cell responses in PWH, and may indicate that PWH have impaired capacity to mount a protective CD8+ T cell response upon SARS-CoV-2 re-infection, particularly with heterologous variants with immune-evading mutations in the spike protein. It is also possible that PWH have expansion of other antigen-specific CD8+ T cells (e.g., CMV-specific) thereby diluting the SARS-CoV-2-specific pool as the denominator was total non-naive CD8+ T cells. Second, we found that SARS-CoV-2 specific CD4+ T cells in PWH had higher expression of the co-inhibitory receptor PD-1, suggesting they may have impaired functionality upon re-encountering infection. Alternatively, PWH may have more SARS-CoV-2 antigen exposure leading to a more exhausted phenotype. Third, we found that a higher CD4/CD8 ratio - an immune marker which can sometimes be optimized with early initiation of ART [39] - was associated with lower expression of PD-1 on SARS-CoV-2-specific CD8+ T cells.

This study has limitations. The sample size was small and the study was underpowered to make comparisons within PWH. Given the nature of the study recruitment, the high prevalence of PASC in our cohort should not be considered to represent the population-level prevalence of PASC, which is likely much lower [40]. In addition, data on other potential confounding factors such as comorbid mental health and substance use were unavailable. Regarding laboratory measurements, IFNγ expression to identify SARS-CoV-2-specific T cells may underestimate the magnitude of the total virus-specific response. Inflammatory marker data was not available on all participants, although sample availability was based on the timing of collection and is not expected to bias the results. Finally, our population of PWH was mostly male, stable on ART, and had strong immune reconstitution, and care should be taken when extrapolating to populations with less access or adherence to ART or those with advanced HIV.

In summary, our analysis provides compelling preliminary evidence suggesting an urgent need to better understand the epidemiology and pathophysiology of PASC within PWH. Such efforts may lead to targeted interventions to prevent or treat this condition among this special population of interest.

## Data Availability

All data produced in the present study are available upon reasonable request to the authors.

## FOOTNOTES

## Acknowledgements

We are grateful to the study participants. We acknowledge current and former LIINC clinical study team members Tamara Abualhsan, Andrea Alvarez, Melissa Buitrago, Monika Deswal, Emily Fehrman, Heather Hartig, Yanel Hernandez, Marian Kerbleski, James Lombardo, Lynn Ngo, Antonio Rodriguez, Dylan Ryder, Ruth Diaz Sanchez, Viva Tai, Cassandra Thanh, Fatima Ticas, Leonel Torres, and Meghann Williams; and LIINC laboratory team members Amanda Buck, Joanna Donatelli, Jill Hakim, Nikita Iyer, Owen Janson, Christopher Nixon, Isaac Thomas, and Keirstinne Turcios. We thank the UCSF AIDS Specimen Bank for processing specimens and maintaining the LIINC biospecimen repository. The intracellular cytokine staining assays were performed in the UCSF Core Immunology Laboratory. We are grateful to Jon Oskarsson, Mary Shiels, Erin Pederson, Parousha Zand, and the UCSF Ward 86 and 360 Positive Health Practices for referring PWH who had COVID-19.

## Author Contributions

MJP, JDK, JNM, SGD, and TJH designed the cohort, which was managed by RH and overseen by MJP. MJP, JDK, SL, RH, JNM, and SGD designed the study instruments. MJP, SEM, AFT, MIA, VT, and RH recruited participants, coordinated research visits, administered study questionnaires, and collected clinical data. CAF, SM, AFT, SAG, JYC, and EOM performed data entry and quality control and addressed or adjudicated issues related to data integrity. SL cleaned the data, maintained the database, and oversaw data management. CY performed the antibody measurements in the laboratory of KLL. NK, RLR, and TJH performed and/or oversaw the cellular immunology assays in the UCSF Core Immunology Laboratory. AC, JWW, and CJP performed and oversaw the inflammatory marker assays at Monogram Biosciences. DW and AS provided the SARS-CoV-2 peptide megapools. MJP, MAS, TMD, SEM, DVG, RLR, and TJH performed the analyses. MJP MAS, TMD, CAF, SEM, SM, SGD, RLR, and TJH drafted the manuscript with input from MSD, PYH, PWH, JFK, JNM, DVG, and MG providing in depth critical review. The study was funded by grants from MAS, MG, SGD, and TJH. All authors reviewed, edited, and approved the manuscript.

