## Supplementary Figure 1 for "POST-ACUTE SEQUELAE AND ADAPTIVE IMMUNE RESPONSES IN PEOPLE LIVING WITH HIV RECOVERING FROM SARS-COV-2 INFECTION"

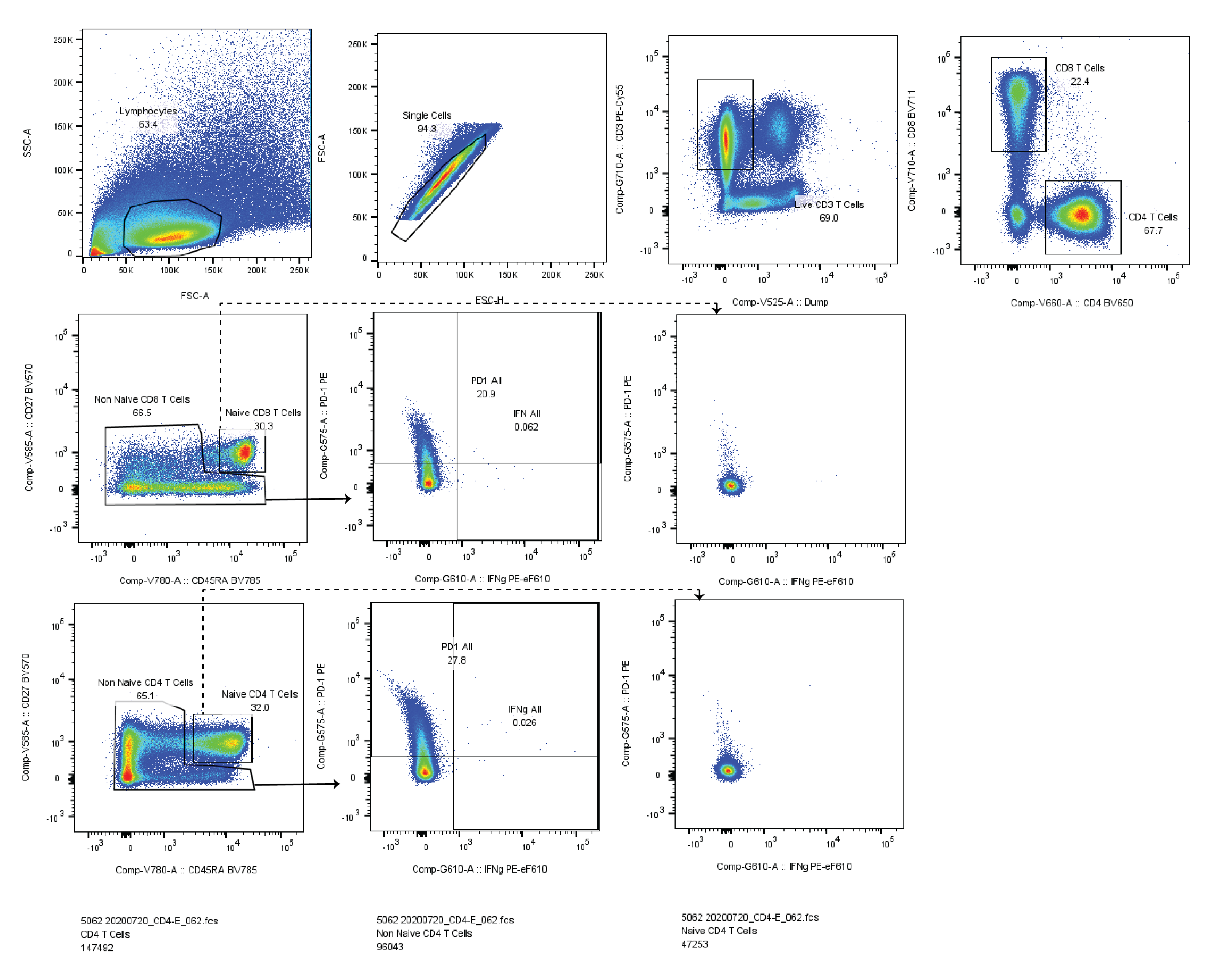

**Supplementary Figure 1**. Example of flow cytometric gating of the intracellular cytokine staining assay in a samples following CD4-optimized SARS-CoV-2 peptide pool stimulation. Percentages of IFNγ CD4 and CD8 T cells and IFNγ positive cell expressing PD-1 were measured for non-naïve (memory) T cells. PD-1 ad IFNγ gating were based on FMOs and by examining expression on naïve T cells (shown to the right denoted by dashed arrows).
