## Supplementary Methods for "POST-ACUTE SEQUELAE AND ADAPTIVE IMMUNE RESPONSES IN PEOPLE LIVING WITH HIV RECOVERING FROM SARS-COV-2 INFECTION"

**SUPPLEMENTAL METHODS**

*Additional information regarding intracellular cytokine staining*

After thawing, PBMCs were rested overnight at 37°C at a concentration of 1x10^6^ PBMC per well in RPMI plus 10% FBS, penicillin, streptomycin, and L-glutamine (R10). The following day, PBMCs were cultured for 18 hours at 37°C in 96-well U-bottom plates at 1x10^6^ PBMC per well in R10 in the presence of either SARS-CoV-2 peptide megapools (described in [[28]](https://paperpile.com/c/GlFoUp/9Kwl); [1 µg/ml/peptide]), or 0.25-1x10^6^ PBMC per well with CD3/CD28 beads (Immunocult, STEMCELL Technologies) as a positive control or equimolar DMSO as negative control. All conditions were in the presence of brefeldin A (Thermo Fisher), monensin (Thermo Fisher) and anti-CD107a. After an 18-hour incubation, cells were washed and surface markers were stained for 15 min at room temp in the dark. Following surface staining, cells were washed twice with PBS and then fixed/permeabilized (BD Cytofix/Cytoperm) for 20 min at 4°C in the dark. Cells were then washed twice with perm wash buffer (BD Perm/Wash) and stained with intracellular antibodies for 30min at 4°C in the dark.
