## Supplementary Table 1 for "POST-ACUTE SEQUELAE AND ADAPTIVE IMMUNE RESPONSES IN PEOPLE LIVING WITH HIV RECOVERING FROM SARS-COV-2 INFECTION"

| **Antigen** | **Channel** | **Clone** | **Vendor** | **Catalogue #** |
| --- | --- | --- | --- | --- |
| CD3 | PE-Cy5.5 | SK7 | ThermoFisher | 35-0036-42 |
| CD4 | BV650 | OKT4 | BioLegend | 317436 |
| CD8a | BV711 | SK1 | BioLegend | 344734 |
| CD14 | BV510 | M5E2 | BioLegend | 301842 |
| CD19 | BV510 | HIB19 | BioLegend | 302242 |
| TCR g/d | BV510 | B1 | BioLegend | 331220 |
| L/D | Aqua | Inv | ThermoFisher | L34966 |
| CD27 | BV570 | O323 | BioLegend | 302825 |
| CD45RA | BV785 | HI100 | BioLegend | 304140 |
| IL-2 | BV421 | MQ1-17H12 | BioLegend | 500328 |
| Granzyme B | FITC | GB11 | BioLegend | 515403 |
| CD107a | AF647 | H4A3 | BioLegend | 328612 |
| TNFa A700 | AF700 | MAb11 | BD Pharmingen | 557996 |
| Perforin | PE-Cy7 | B-D48 | BioLegend | 353316 |
| IFNg | PE-CF594 | B27 | BD Pharmingen | 562392 |
| PD-1 | PE | EH12.2H7 | BioLegend | 329906 |

**Supplemental Table 1.** Key reagents and resources.
